# Muscle Strength and Muscle Mass as Predictors of Hospital Length of Stay in Patients with Moderate to Severe COVID-19: A Prospective Observational Study

**DOI:** 10.1101/2021.03.30.21254578

**Authors:** Saulo Gil, Wilson Jacob Filho, Samuel Katsuyuki Shinjo, Eduardo Ferriolli, Alexandre Leopold Busse, Thiago Junqueira Avelino-Silva, Igor Longobardi, Gersiel Nascimento de Oliveira, Paul Swinton, Bruno Gualano, Hamilton Roschel, on behalf of the HCFMUSP COVID-19 Study Group, Eloisa Bonfá, Edivaldo Utiyama, Aluisio Segurado, Beatriz Perondi, Anna Miethke Morais, Amanda Montal, Leila Letaif, Solange Fusco, Marjorie Fregonesi Rodrigues da Silva, Marcelo Rocha, Izabel Marcilio, Izabel Cristina Rios, Fabiane Yumi Ogihara Kawano, Maria Amélia de Jesus, Ésper Georges Kallas, Carolina Carmo, Clarice Tanaka, Heraldo Possolo de Souza, Julio F. M. Marchini, Carlos Carvalho, Juliana Carvalho Ferreira, Anna Sara Shafferman Levin, Maura Salaroli de Oliveira, Thaís Guimarães, Carolina dos Santos Lázari, Alberto José da Silva Duarte, Ester Sabino, Marcello Mihailenko Chaves Magri, Tarcisio E. P. Barros-Filho, Maria Cristina Peres Braido Francisco

**Affiliations:** Applied Physiology & Nutrition Research Group, School of Physical Education and Sport, Rheumatology Division, Faculdade de Medicina FMUSP, Universidade de Sao Paulo, Av. Prof. Mello Moraes, 65 - São Paulo, SP, Brazil; Rheumatology Division, Hospital das Clinicas HCFMUSP, Faculdade de Medicina FMUSP, Universidade de Sao Paulo, Sao Paulo, SP, BR; Laboratorio de Investigacao Medica em Envelhecimento (LIM-66), Servico de Geriatria, Hospital das Clinicas HCFMUSP, Faculdade de Medicina, Universidade de Sao Paulo, Sao Paulo, SP, BR; Division of Internal and Geriatric Medicine, Department of Internal Medicine – Ribeirão Preto Medical School, Universidade de Sao Paulo, Ribeirao Preto, SP, BR; School of Health Sciences, Robert Gordon University, Garthdee Road, Aberdeen, UK

## Abstract

**Importance:** Strength and muscle mass are predictors of relevant clinical outcomes in critically ill patients, but in hospitalized patients with COVID-19 remains to be determined.

**Objective:** To investigate whether muscle strength or muscle mass are predictive of hospital length of stay (LOS) in patients with moderate to severe COVID-19.

**Design:** Prospective observational study.

**Setting:** Clinical Hospital of the School of Medicine of the University of Sao Paulo.

**Participants:** One hundred ninety-six patients were evaluated. Ten patients did not test positive for SARS-CoV-2 during hospitalization and were excluded from the analyses. The sample comprised patients of both sexes (50% male) with a mean age (SD) of 59 (±15) years, body mass index of 29.5 (±6.9) kg/m^2^. The prevalence of current smoking patients was 24.7%, and more prevalent coexisting conditions were hypertension (67.7%), obesity (40.9%), and type 2 diabetes (36.0%). Mean (SD) LOS was 8.6 days (7.7); 17.0% of the patients required intensive care; 3.8% used invasive mechanical ventilation; and 6.6% died during the hospitalization period.

**Main outcome:** The outcome was LOS, defined as time from hospital admission to medical discharge.

**Results:** The crude Hazard Ratio (HR) for LOS was greatest for handgrip strength comparing the strongest *vs*. other patients (1.54 [95%CI: 1.12 – 2.12; p = 0.008]). Evidence of an association between increased handgrip strength and shorter hospital stay was also identified when handgrip strength was standardized according to the sex-specific mean and standard deviation (1.23 [95%CI: 1.06 – 1.19; p = 0.008]). The magnitude of these associations remained consistent and statistically significant after adjusting for other covariates. Mean LOS was shorter for the strongest patients (7.5 ± 6.1 days) *vs*. others (9.2 ± 8.4 days). Evidence of associations were also present for vastus lateralis cross-sectional area. The crude HR identified shorter hospital stay for patients with greater sex-specific standardized values (1.17 [95%CI: 1.01 – 1.36; p = 0.037]); however, we found increased uncertainty in the estimate with the addition of other covariates (1.18 [95%CI: 0.97 – 1.43; p = 0.092]). Evidence was also obtained associating longer hospital stays for patients with the lowest values for vastus lateralis cross-sectional area (0.69 [95%CI: 0.50 – 0.95; p = 0.025). Mean LOS for the patients with the lowest muscle cross-sectional area was longer (10.8 ± 8.8 days) *vs*. others (7.7 ± 7.2 days).

**Conclusions and Relevance:** Muscle strength and mass assessed upon hospital admission are predictors of LOS in patients with moderate to severe COVID-19, which stresses the value of muscle health in prognosis of this disease.

**Funding:** The authors acknowledge the support by the Brazilian National Council for Scientific and Technological Development (CNPq - grant 301571/2017-1). H.R. and B.G. are supported by grants from the Conselho Nacional de Pesquisa e Desenvolvimento (CNPq 428242/2018-9; 301571/2017-1; 301914/2017-6). B.G. is also supported by a grant from the Sao Paulo Research Foundation (FAPESP 2017/13552-2).

**Key points:** *Question:* Do muscle strength and muscle mass predict hospital length of stay (LOS) in patients with moderate to severe COVID-19 patients?

*Findings:* In this prospective observational study that included 186 hospitalized patients with moderate to severe COVID-19, we observed that LOS was shorter among patients in the highest tertile of strength (assessed by handgrip) *vs*. those in the mid/lowest tertiles (crude Hazard Ratio [HR]: 1.54, 95%CI: 1.12-2.12). In addition, LOS was longer among patients in the lowest tertile of muscle cross-sectional area (assessed by ultrasound imaging) *vs*. those in the mid/highest tertiles (HR: 0.69, 95%CI: 0.50 – 0.95).

*Meaning:* Muscle strength and mass assessed on hospital admission are predictors of LOS in patients with moderate to severe COVID-19, suggesting that muscle health may be protective in this disease.

## Introduction

Aging and chronic conditions such as type 2 diabetes increase the risk of developing severe forms of COVID-19. Nevertheless, apparently healthier, younger individuals may also require hospitalization and develop poor outcomes ^1-3^. This suggests that there might be undiscovered clinical features associated with COVID-19 prognosis, with muscular parameters being potential candidates.

Skeletal muscle constitutes ∼40% of total body mass and plays a pivotal role in different physiological process such as immune response, regulation of glucose levels, protein synthesis, and basal metabolic rate ^4-6^. Handgrip strength and muscle mass have been shown previously to be predictive of clinical outcomes, such as hospital length of stay (LOS) and mortality, in distinct populations ^7-10^. The prognostic value of skeletal muscle parameters among hospitalized patients with COVID-19 remains to be determined. Herein, we investigated whether muscle strength and muscle mass assessed at hospital admission are predictive of LOS in patients with moderate to severe COVID-19

## Methods

### Study design

This is a prospective observational study conducted between March 2020 and October 2020 in the Clinical Hospital of the School of Medicine of the University of Sao Paulo in Brazil (HCFMUSP) the largest quaternary referral teaching hospital in Latin America. This study was approved by the local Ethics Committee (Ethics Committee Approval Number (31303720.7.0000.0068). All patients provided written informed consent before entering the study. This manuscript was reported according to the Strengthening the Reporting of Observational Studies in Epidemiology (STROBE) Statement.

### Participants

The inclusion criteria were: 1) aged 18 or older; 2) diagnosis of COVID-19 by PCR for SARS-CoV-2 from nasopharyngeal swabs or computed tomography scan findings (bilateral multifocal ground-glass opacities ≥ 50%) compatible with the disease; 3) diagnosis of flu syndrome with hospitalization criteria on hospital admission, presenting respiratory rate ≥ 24 breaths per minute, saturation < 93% on room air or risk factors for complications, such as heart disease, diabetes mellitus, systemic arterial hypertension, neoplasms, immunosuppression, pulmonary tuberculosis, and obesity, followed by COVID-19 confirmation. Exclusion criteria were: 1) cancer in the past 5 years; 2) delirium; 3) cognitive deficit that precluded the patient from reading and signing the informed consent form; 4) prior diagnosis of muscle degenerative disease (e.g., myopathies, amyotrophic lateral sclerosis, stroke); 5) patients already admitted under invasive mechanical ventilation. Patients who met these criteria were considered to have moderate to severe COVID-19.

### Data collection

All patients were evaluated in the point-of-care within less than 48 hours upon hospital admission for handgrip strength and vastus lateralis cross-sectional area, by means of ultrasound imaging, and were followed until medical discharge.

Handgrip strength assessments were performed with the patient seated holding the dynamometer (TKK 5101; Takei, Tokyo, Japan) with the dominant hand and elbow positioned at a 90° angle. Three maximum attempts of 5 seconds with 1 minute of the interval between attempts were performed and the best result was used for analysis.

Vastus lateralis cross-sectional area was assessed by a B-mode ultrasound with a 7.5-MHz linear-array probe (SonoAce R3, Samsung-Medison, Gangwon-do, South Korea) as previously described ^11^. Cross-sectional area analyses were performed in a blinded fashion by a single investigator using ImageJ (NIH, USA). Demographic, clinical, and biochemical data of the patients were obtained through medical records.

### Outcome and stratification of patients

Our primary outcome was LOS, defined as time (days) from hospital admission to medical discharge. Patients who died during the hospitalization period were right-censored in the LOS analysis. To examine whether muscle strength or mass were predictive of LOS, we ranked patients according to handgrip strength and vastus lateralis cross-sectional area into sex-specific tertiles. Then, we compared the highest tertile (High) *vs*. the combined mid and lowest tertiles (High *vs*. Other), and the lowest tertile (Low) *vs*. the combined mid and highest tertiles (Low *vs*. Other)

### Sample size and statistical analyses

An a-priori sample size estimate was made to achieve small optimism in the predictor effect estimates as defined by a global shrinkage factor of 0.9 ^12^. The expected shrinkage is conditioned on the sample size (n), the total number of predictors (p) and a generalization of the proportion of variance explained for multivariable models with time-to-event outcomes 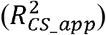 ^12^. For the present study p was set to 6 and 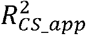 to 0.5 based on findings from previous research ^7, 13, 14^ and therein, indicating a required sample size of n = 184. Guided by this estimate, a total of 196 patients were evaluated during their hospital stay.

The outcome (LOS) was analyzed with multivariable Cox proportional baseline hazard models and adjusted for sex (male or female), age group (18-35, 36-55 or ≥ 56), obesity (BMI<30 or BMI≥30), oxygen support at admission (0-4L, 5-9L, ≥10L) and Type 2 diabetes (yes or no). Both crude and adjusted hazard ratios (HRs) were estimated for handgrip strength and vastus lateralis cross-sectional area. Each predictor was assessed as both a discrete and continuous predictor. Discrete models were conducted by calculating sex-specific tertiles and then focusing on either the largest tertile (High *vs*. Other) or the smallest tertile (Low *vs*. Other). Continuous models were conducted by standardizing predictor values relative to the sex-specific mean and standard deviation. HRs were accompanied with corresponding 95% confidence intervals (95%IC), with all analyses performed in the statistical environment R (version 3.5.3; R Core Team 2020) with the survival ^15^ and survminer ^16^ packages.

## Results

### Patients

One hundred ninety-six patients were evaluated. Ten patients did not test positive for SARS-CoV-2 during the hospitalization period and were excluded from the analyses. Table 1 shows the demographic, biochemical, and clinical characteristics of the patients. Overall, 86% (160 of 186) had a positive PCR test for SARS-CoV-2 at the enrollment, and 42% had computed tomography scan findings suggestive (i.e., pulmonary commitment ≥50%) for COVID-19. All the remaining patients (26 of 186) had the diagnosis confirmed by serology assay to detect IgG against SARS-CoV-2 at some point during the hospital day. The sample comprised patients of both sexes (50% male) with a mean (SD) age of 59 years (±15), body mass index of 29.5 kg/m^2^ (±6.9). The prevalence of current smoking patients was 24.7%, and more prevalent coexisting conditions were hypertension (67.7%), obesity (40.9%), and type 2 diabetes (36.0%).

**Table 1.**
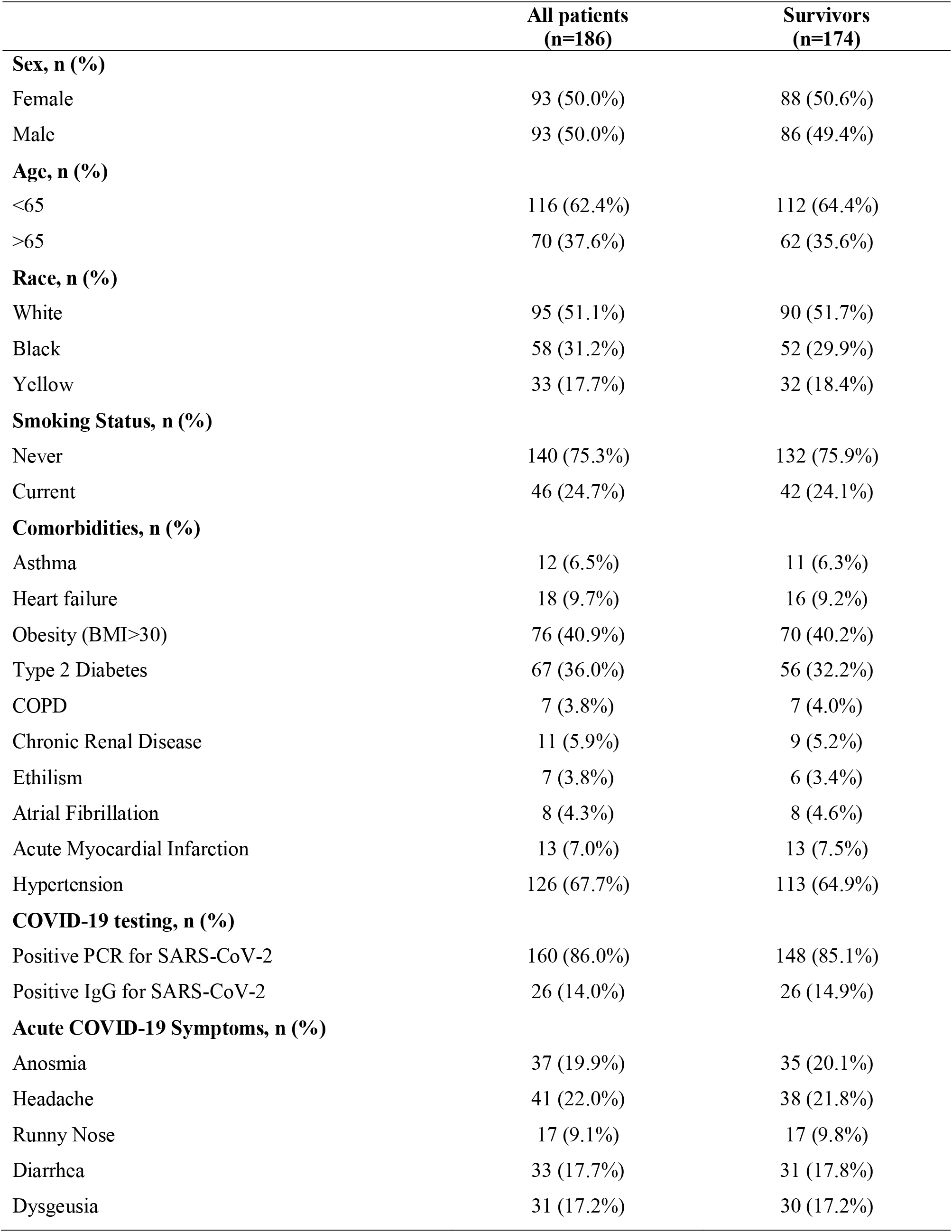
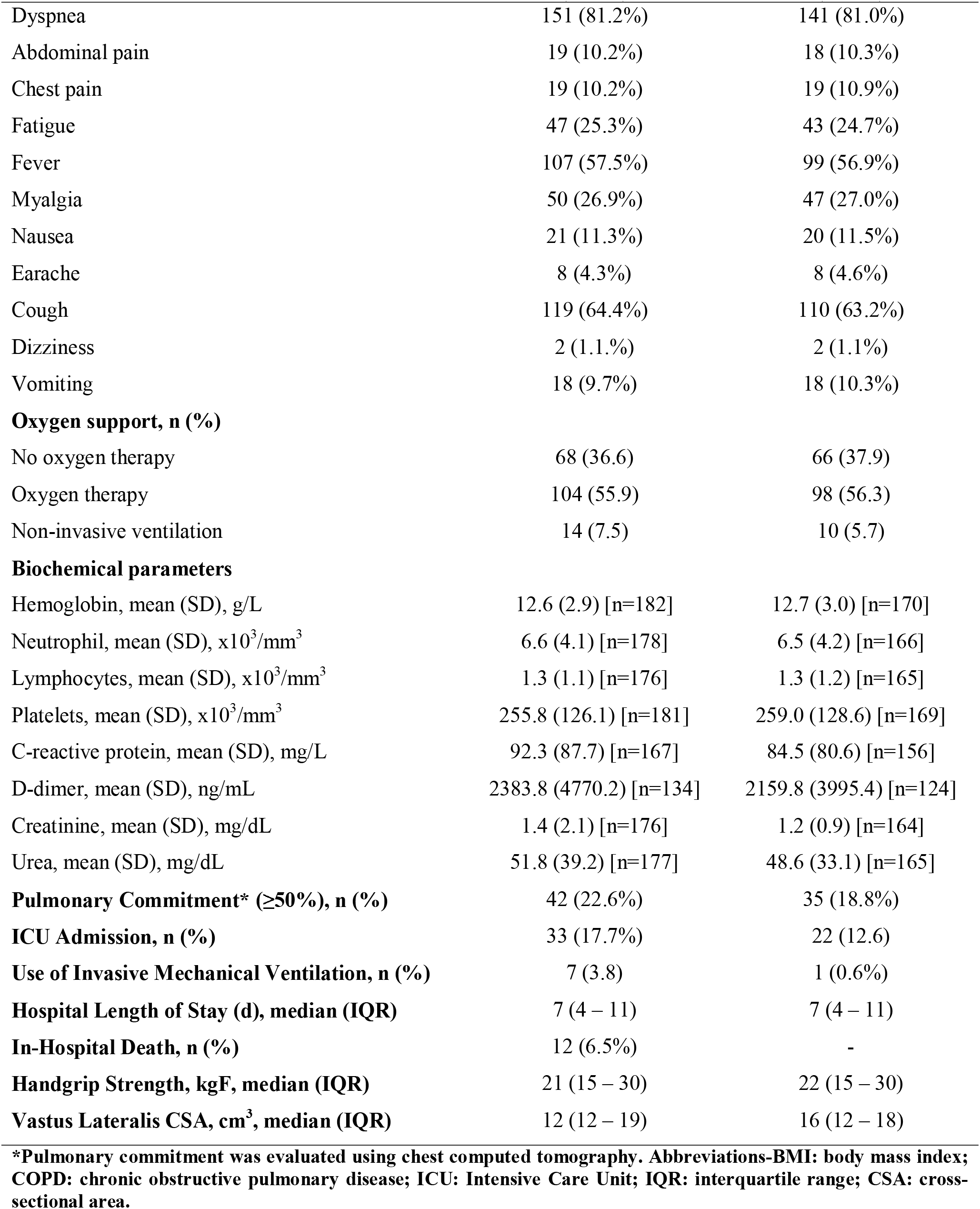
Demographics and clinical characteristics of patients at hospital admission.

The signs and symptoms more commonly observed at admission were dyspnea (81.2%), cough (64.4%), fever (57.5%), myalgia (26.9%), fatigue (25.3%), headache (22.0%), anosmia (19.9%), diarrhea (17.7%), dysgeusia (17.2%), nausea (11.3%), abdomen pain (10.2%), chest pain (10.2%), vomiting (9.7%), runny nose (9.1%), earache (4.3%), and dizziness (1.1%). Mean (SD) LOS was 8.6 days (7.7); 17.0% of the patients required intensive care; 3.8% used invasive mechanical ventilation; and 6.6% died during the hospitalization period.

### Primary Outcome

The crude HR for time from hospital admission to discharge was greatest for handgrip strength comparing the strongest *vs*. other patients (1.54 [95%CI: 1.12 – 2.12; p = 0.008]). Evidence of an association between increased handgrip strength and shorter hospital stay was also identified when handgrip strength was standardized according to the sex-specific mean and standard deviation (1.23 [95%CI: 1.06 – 1.19; p = 0.008).

The magnitude of these associations remained consistent and statistically significant after adjusting for other covariates (Table 2). Mean LOS was shorter for the strongest patients (7.5 ± 6.1 days) *vs*. others (9.2 ± 8.4 days).

**Table 2.**
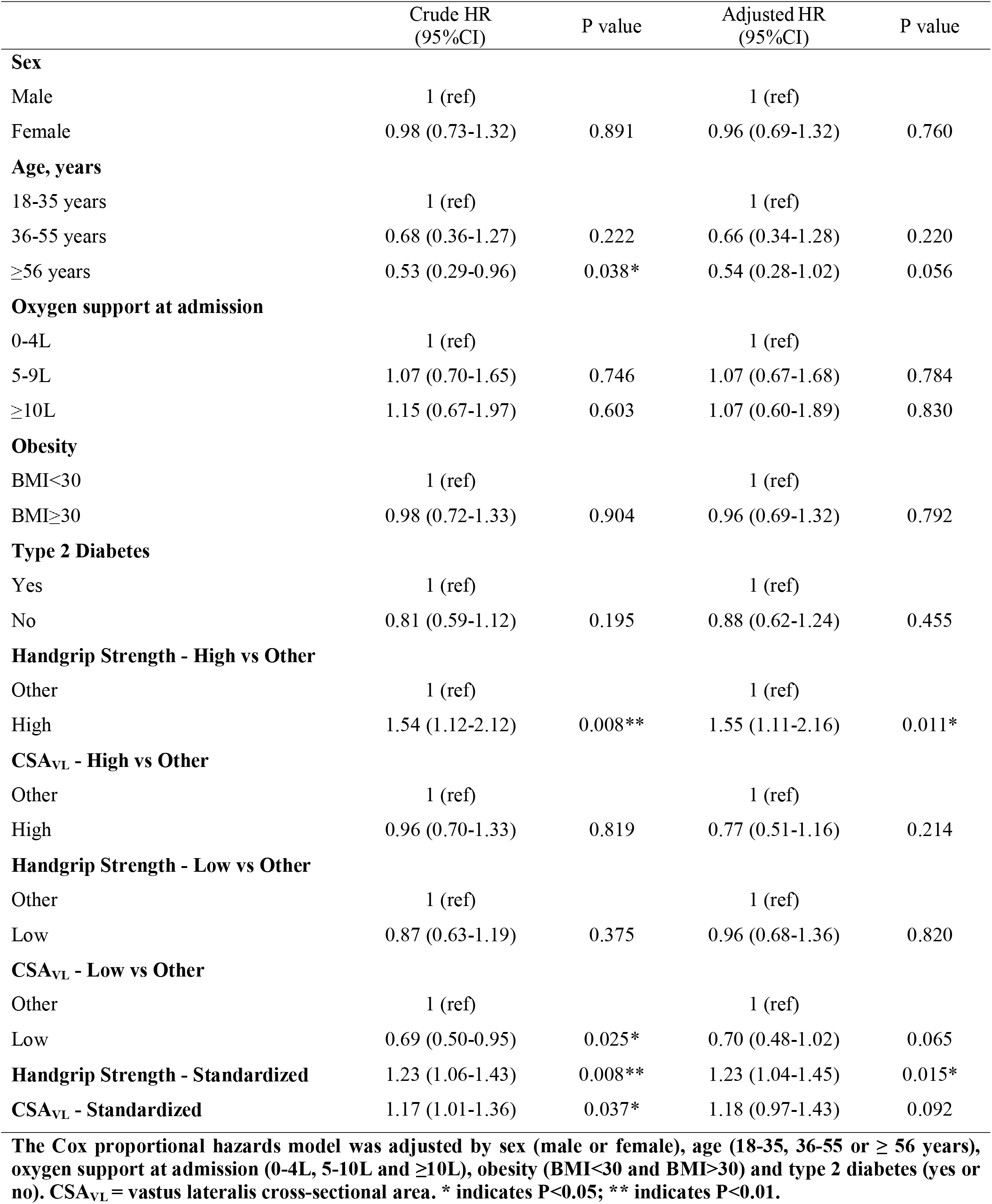
Crude and adjusted hazard Ratio (HR) for hospital length of stay.

Evidence of associations were also present for vastus lateralis cross-sectional area. The crude HR identified shorter hospital stay for patients with greater sex-specific standardized values (1.17 [95%CI: 1.01 – 1.36; p = 0.037); however, we found increased uncertainty in the estimate with the addition of other covariates (1.18 [95%CI: 0.97 – 1.43; p = 0.092). Evidence was also obtained associating longer hospital stays for patients with the lowest values for vastus lateralis cross-sectional area (0.69 [95%CI: 0.50 – 0.95; p = 0.025). Mean LOS for the patients with the lowest muscle cross-sectional area was longer (10.8 ± 8.8 days) *vs*. others (7.7 ± 7.2 days).

## Discussion

In this prospective observational study, we found muscle strength (as assessed by handgrip) and muscle mass (as assessed by vastus lateralis cross-sectional area) are predictive of LOS in hospitalized patients with moderate to severe COVID-19. To the best of our knowledge, this is the first study to demonstrate the prognostic value of these skeletal muscle parameters in this disease.

A recent study demonstrated that the Clinical Frailty Score (CFS) independently predicted time to medical discharge and mortality in COVID-19 patients ^13^. Despite the value of these findings, it is noteworthy that the CFS is a judgement-based frailty tool that relies highly on experience and training for proper categorization of the patients. Moreover, CFS is ultimately an indirect measure of functional status and is mainly used in geriatric patients. These are factors that might limit the reliability of CFS in real-life clinical scenarios. Conversely, handgrip strength is a simple, direct, easy handling, low-cost measurement commonly utilized in the clinical setting as an indicator of the general health status in individuals across a wide age range. Indeed, handgrip strength assessed at hospital admission have been shown to be a predictive measure of LOS and mortality in distinct populations ^7-10^. Our findings extend this knowledge to patients admitted into the hospital with acute COVID-19 symptoms, by showing that stronger patients had lower LOS than their weaker counterparts.

Muscle mass is also considered as an indicator of general health status ^17, 18^. Previous studies have suggested that low muscle mass (assessed by mid-arm circumference, calf circumference, and estimated by anthropometric equations) may predict mortality among elderly ^17, 18^. In the current study, we directly assessed, in the point-of-care, vastus lateralis cross-sectional area using ultrasonography among patients with COVID-19. Our findings suggest that low muscle mass could contribute to higher LOS among COVID-19 patients During a critical illness, net breakdown of muscle protein is stimulated to provide abundant amino acids to meet these increased demands of tissues such as immune cells and liver ^19^. In this context, patients with limited muscle mass reserves would presumably be more vulnerable to stress factors, such as severe burn injuries and cancer ^20, 21^. The present findings suggest that this could be the case of COVID-19.

Muscle mass plays a key role in recovery from critical illness, whereas muscle strength and function are key to the recovery process ^22^. If there is a preexisting deficiency of muscle mass before the onset of an acute illness, one may speculate that the expected loss of muscle mass and function associated with hospitalization may push the patient over a threshold that makes recovery of normal function unlikely to ever occur ^22^. The impact of this physiopathological mechanism on long-term effects of COVID-19 (long COVID) remains to be explored.

### Limitations

First, the longitudinal design of this study does not allow causative conclusions. Second, although this study was adequately powered to detect changes in the selected outcomes, this was still a small cohort composed by patients with heterogeneous clinical features, medication regimen and disease manifestations, possibly subject to unmeasured confounders. While the Cox proportional hazards models were controlled for several potentially confound variables, direct sub-group comparisons were not possible due to sample size constraints. Third, our results are confined to patients with moderate to severe COVID-19 and should be read with care regarding other clinical settings. Finally, the minimal clinically important difference in LOS among patient with COVID-19 is yet unknown, which limits the ability to make clinical inferences about the present findings.

### Conclusions

Muscle strength and mass assessed on hospital admission are predictors of LOS in patients with COVID-19. While it is unknown whether these muscular parameters add to the prognostic value provided by the more established and accepted predictors that already have been identified ^23^, the present data suggest that muscle health may benefit patients with moderate to severe COVID-19. The evidence provided by this study paves the way for randomized controlled trials to test the utility of preventive or in-hospital interventions in shortening LOS among these patients through improving muscle mass and/or function.

## Data Availability

Further information and requests for database should be directed to and will be fulfilled by the Lead Contact, Hamilton Roschel.

## Acknowledgements

The authors are thankful to the task force of HCFMUSP COVID-19 Study Group: Rosemeire Keiko, Danielle Pedroni de Moraes, Renato Madrid Baldassare, Antônio José Pereira, Elizabeth de Faria, Gisele Pereira, Lucila Pedroso da Cruz, Marcelo, Cristiano de Azevedo Ramos, Vilson Cobello Junior.

## Figure Legends

**Figure 1.**
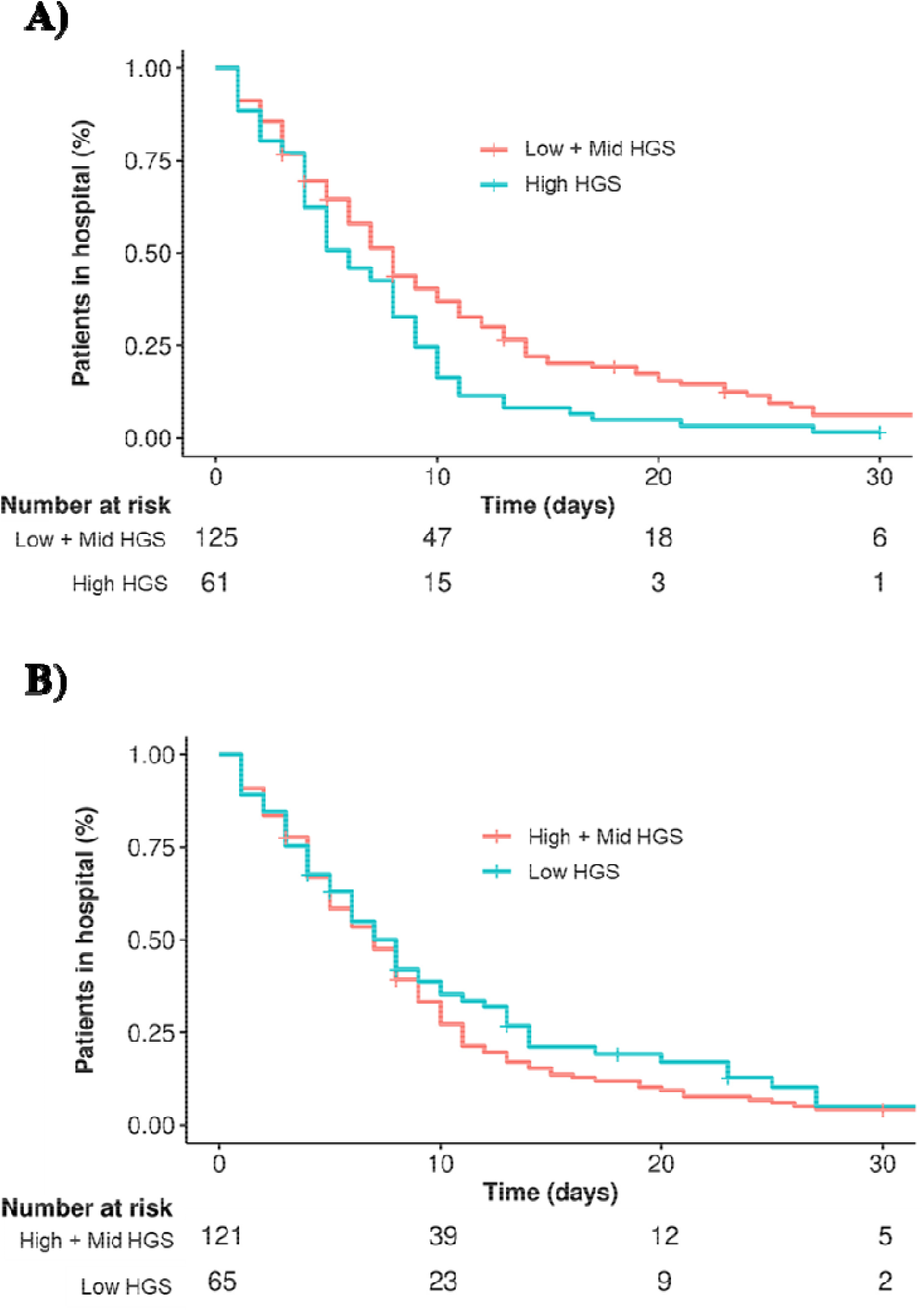
Kaplan-Meier plot of time from hospital admission to hospital discharge according to handgrip strength.

**Figure 1.**
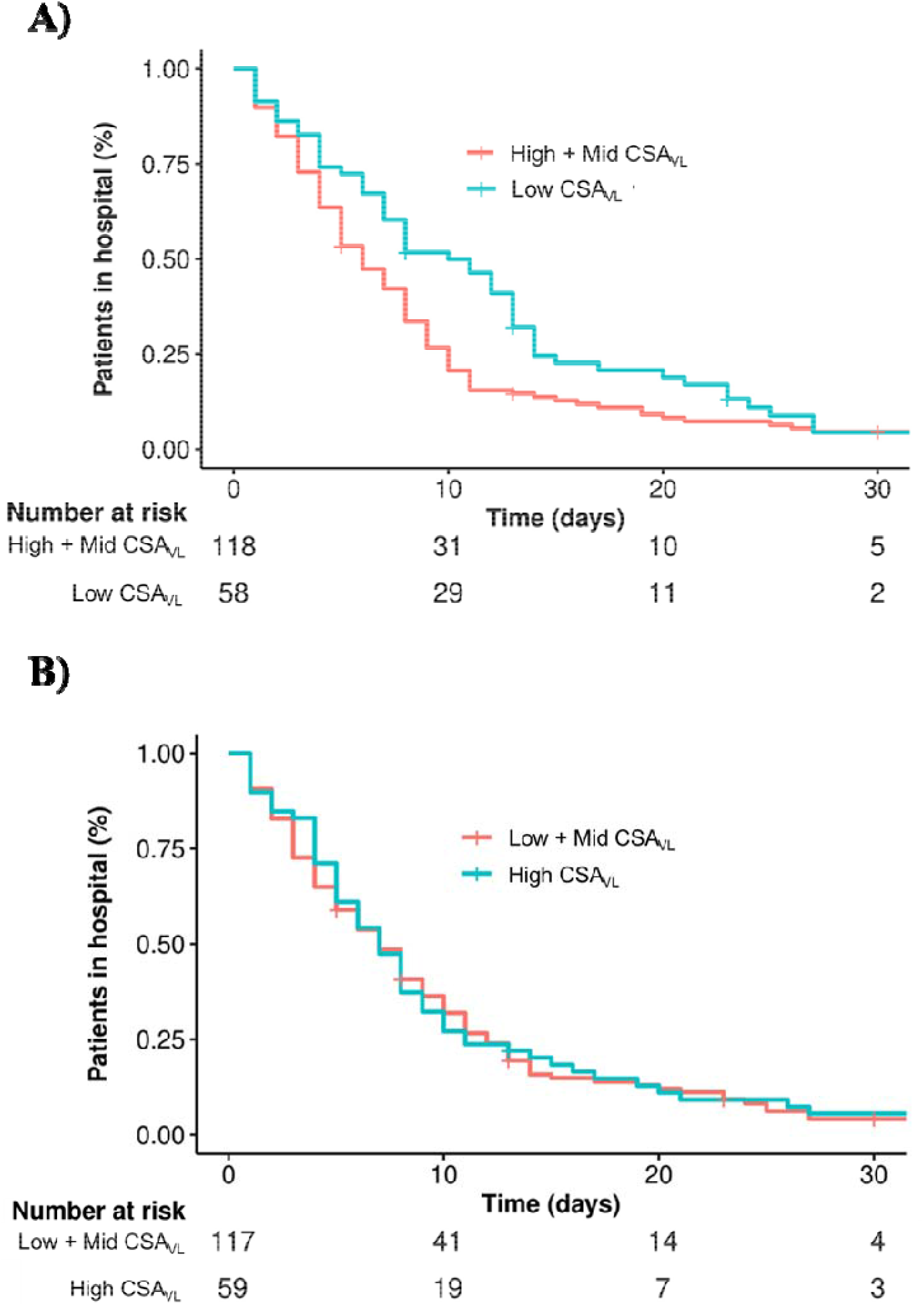
Kaplan-Meier plot of time from hospital admission to hospital discharge according to vastus lateralis cross-sectional area.

## References

1. Guan WJ, Ni ZY, Hu Y, et al. Clinical Characteristics of Coronavirus Disease 2019 in China. N Engl J Med. Apr 30 2020;382(18):1708–1720. doi:10.1056/NEJMoa2002032

2. Huang C, Wang Y, Li X, et al. Clinical features of patients infected with 2019 novel coronavirus in Wuhan, China. Lancet. Feb 15 2020;395(10223):497–506. doi:10.1016/S0140-6736(20)30183-5

3. Richardson S, Hirsch JS, Narasimhan M, et al. Presenting Characteristics, Comorbidities, and Outcomes Among 5700 Patients Hospitalized With COVID-19 in the New York City Area. JAMA. May 26 2020;323(20):2052–2059. doi:10.1001/jama.2020.6775

4. Brandt C, Pedersen BK. The role of exercise-induced myokines in muscle homeostasis and the defense against chronic diseases. J Biomed Biotechnol. 2010;2010:520258. doi:10.1155/2010/520258

5. Egan B, Zierath JR. Exercise metabolism and the molecular regulation of skeletal muscle adaptation. Cell metabolism. Feb 5 2013;17(2):162–84. doi:10.1016/j.cmet.2012.12.012

6. Lightfoot A, McArdle A, Griffiths RD. Muscle in defense. Crit Care Med. Oct 2009;37(10 Suppl):S384–90. doi:10.1097/CCM.0b013e3181b6f8a5

7. Mendes J, Azevedo A, Amaral TF. Handgrip strength at admission and time to discharge in medical and surgical inpatients. JPEN Journal of parenteral and enteral nutrition. May 2014;38(4):481–8. doi:10.1177/0148607113486007

8. Moisey LL, Mourtzakis M, Cotton BA, et al. Skeletal muscle predicts ventilator-free days, ICU-free days, and mortality in elderly ICU patients. Crit Care. Sep 19 2013;17(5):R206. doi:10.1186/cc12901

9. Mueller N, Murthy S, Tainter CR, et al. Can Sarcopenia Quantified by Ultrasound of the Rectus Femoris Muscle Predict Adverse Outcome of Surgical Intensive Care Unit Patients as well as Frailty? A Prospective, Observational Cohort Study. Annals of surgery. Dec 2016;264(6):1116–1124. doi:10.1097/SLA.0000000000001546

10. Weir DE, Tingley J, Elder GC. Acute passive stretching alters the mechanical properties of human plantar flexors and the optimal angle for maximal voluntary contraction. Eur J Appl Physiol. Mar 2005;93(5-6):614–23.

11. Lixandrao ME, Damas F, Chacon-Mikahil MP, et al. Time Course of Resistance Training-Induced Muscle Hypertrophy in the Elderly. J Strength Cond Res. Jan 2016;30(1):159–63. doi:10.1519/JSC.0000000000001019

12. Riley RD, Snell KI, Ensor J, et al. Minimum sample size for developing a multivariable prediction model: PART II -binary and time-to-event outcomes. Stat Med. Mar 30 2019;38(7):1276–1296. doi:10.1002/sim.7992

13. Hewitt J, Carter B, Vilches-Moraga A, et al. The effect of frailty on survival in patients with COVID-19 (COPE): a multicentre, European, observational cohort study. The Lancet Public health. Aug 2020;5(8):e444–e451. doi:10.1016/S2468-2667(20)30146-8

14. Kerr A, Syddall HE, Cooper C, Turner GF, Briggs RS, Sayer AA. Does admission grip strength predict length of stay in hospitalised older patients? Age Ageing. Jan 2006;35(1):82–4. doi:10.1093/ageing/afj010

15. Therneau T. A Package for Survival Analysis in R. R Package Version 3.1–12,. Accessed 11 June 2020,

16. Kassambara AK, M.; Biecek P. Survminer: Drawing Survival Curves Using’Ggplot2’. R PackageVersion 0.4.6. Accessed11 June 2020,

17. Wang H, Hai S, Liu Y, Liu Y, Dong B. Skeletal Muscle Mass as a Mortality Predictor among Nonagenarians and Centenarians: A Prospective Cohort Study. Scientific reports. Feb 20 2019;9(1):2420. doi:10.1038/s41598-019-38893-0

18. Weng CH, Tien CP, Li CI, et al. Mid-upper arm circumference, calf circumference and mortality in Chinese long-term care facility residents: a prospective cohort study. BMJ open. May 9 2018;8(5):e020485. doi:10.1136/bmjopen-2017-020485

19. Wolfe RR, Martini WZ. Changes in intermediary metabolism in severe surgical illness. World journal of surgery. Jun 2000;24(6):639–47. doi:10.1007/s002689910105

20. Kadar L, Albertsson M, Areberg J, Landberg T, Mattsson S. The prognostic value of body protein in patients with lung cancer. Ann N Y Acad Sci. May 2000;904:584–91. doi:10.1111/j.1749-6632.2000.tb06520.x

21. Pereira CT, Barrow RE, Sterns AM, et al. Age-dependent differences in survival after severe burns: a unicentric review of 1,674 patients and 179 autopsies over 15 years. Journal of the American College of Surgeons. Mar 2006;202(3):536–48. doi:10.1016/j.jamcollsurg.2005.11.002

22. Wolfe RR. The underappreciated role of muscle in health and disease. Am J Clin Nutr. Sep 2006;84(3):475–82. doi:10.1093/ajcn/84.3.475

23. Wongvibulsin S, Garibaldi BT, Antar AAR, et al. Development of Severe COVID-19 Adaptive Risk Predictor (SCARP), a Calculator to Predict Severe Disease or Death in Hospitalized Patients With COVID-19. Ann Intern Med. Mar 2 2021;doi:10.7326/M20-6754

